# Inclusive Costs of NPI Measures for COVID-19 Pandemic: Three Approaches

**DOI:** 10.1101/2020.03.26.20044552

**Authors:** Alexander Ugarov

## Abstract

The paper evaluates total inclusive costs of three public health approaches to address the COVID-19 epidemic in the US based on epidemiological projections in Ferguson et al (2020). We calculate and add costs of lost productivity and costs of mortality measured through the value of statistical life. We find that the aggressive approach which involves strict suppression measures and a drastic reduction of economic activity for three months with extensive testing and case tracking afterwards results in the lowest total costs for the society. The approach of doing no non-pharmaceutical measures results in the lowest total costs if the infection fatality rate falls below 0.15%.

## 1 Introduction

As more and more countries are imposing strict measures to suppress the COVID-19 pandemic, there is a growing concern that these measures increase total societal costs. Existing literature suggests that mitigation policies do not necessarily provide net benefits to the society (Mesnard and Seabright, 2009). While mitigation decreases deaths, it also reduces economic activity. For example, preliminary data from China suggests approximately 10% decline in GDP in the first quarter of 2020^1^.

This paper conducts the first (to out knowledge) cost-benefit analysis of potential mitigation measures for the COVID-19 epidemic in the US. We calculate total societal costs of the pandemic and corresponding non-pharmaceutical measures. These costs include three components: costs of mortality, costs of illness and costs of reduced economic activity due to mitigation and precautionary demand.

We put a monetary value on COVID-19 deaths by using the value of statistical life approach. While it seems weird to put a monetary estimate on life, people often do it implicitly while deciding to save time by speeding or deciding to save money by not investing in tornado shelters. The value of statistical life represents the amount of money people willingly accept for increasing their own risk of death. This approach becomes more common in public health literature (Jamison et al, 2013) and is already standard for many environmental protection agencies both in the US and abroad.

This cost-benefit analysis calculates monetary estimates of total costs for three different approaches. The **laissez-faire** implies doing nothing. The **herd immunity approach** applies non-pharmaceutical measures to keep the number of new cases at the maximum of healthcare capacity. The third **aggressive approach** uses the most aggressive non-pharmaceutical suppression and mitigation measures to reduce the number of new cases and then tries to eliminate the virus through extensive testing, case tracking and case isolation. We use the results of epidemiological modeling (Ferguson et al, 2020) for the first two approaches and our conservative estimates based on existing data for the third approach to project the number of infections, number of critical cases and the number of deaths. We then calculate costs of mitigation measures and of the disease itself for total output and add these costs to the costs of lives lost estimated through the value of statistical life approach.

We find that the aggressive approach achieves the lowest total costs for the society for any reasonable value of statistical life (VSL) and for baseline epidemiological assumptions. The herd immunity approach dominates the laissez-faire approach of doing nothing for most of the VSL range except for the lowest end. The laissez-faire approach starts to completely dominate the herd immunity approach if the infection fatality rate (IFR) falls below 0.3% and it starts dominating the aggressive approach completely for IFR below 0.15%.

The calculus of total costs would shift more in favor of the herd immunity approach by accounting for increasing capacity of the healthcare system, gradual elimination of suppression measures and for the positive effect of suppression measures on infections from other airborne diseases. On another hand, the herd immunity approach might be completely unfeasible if the immunity from the virus lasts less than 2 years, while the laissez-faire and aggressive approaches might results in a complete eradication of the virus in human population.

## 2 Method and Assumptions

### 2.1 Potential Policies

We consider three potential policy approaches. Two of these approaches are consistent with approaches suggested in Ferguson et al (2020). The aggressive approach is not a part of Ferguson et al (2020), but roughly follows the steps taken by China and several other Asian countries (South Korea, Taiwan, Japan, Singapore). All of these policies are assumed to start in the US at the beginning of April.

#### Laissez-faire Approach

Federal, state and local authorities take no mitigation or suppression measures in this scenario. Population can still take precautions if their personal benefits exceed personal costs. This scenario is closest to the approach used in the 2009 flu pandemic and other flu epidemics with lower mortality rates.

#### Herd Immunity Approach

In the herd immunity approach, the state imposes multiple NPI in order to bring R0 down to the level slightly above one. Then, after the number of infections gets to close to the healthcare capacity and while the proportion of immune individuals in the population grows, it slowly dismantle NPI measures in order to keep the number of new infections at the level of healthcare capacity.

The analysis by Ferguson et al (2020) suggests that the combination of three measures implements this approach: school closures, home quarantines and social distancing of the entire population. Any smaller combination of measures is insufficient for reducing the reproductive number to one and would put an over-stress on the healthcare system. School closures imply that all public schools close and children have to stay at home with their parents. Home quarantines mean that the whole household avoids going outside (including to work) for some period if any household member tests positive for COVID-19. We interpret social distancing to mean closure or complete avoidance of food services and drinking places, entertainment, arts and recreation as consuming these services conflicts with social distancing.

#### Aggressive Approach

In this approach, the government imposes extremely strict measures in order to suppress the epidemic and to bring the number of new cases close to zero. These measures last three months from April to June. The number of new cases is then held at the level around zero without strict movement and distancing restrictions through mass testing, case tracking and isolation.

The experience of China in January-February 2020 demonstrates the the aggressive approach can effectively reduce the number of cases to almost zero. These measure included extensive testing, mask wearing, shutting down most non-essential businesses and strict movement restrictions (shelter-in-place, lock-down policies). The experience of South Korea, Singapore and Taiwan demonstrates that for low number of infections, case tracking and isolation can keep the epidemic at bay without strict movement restrictions.

### 2.2 Epidemiological Assumptions

The disease has 0.9% infection fatality rate meaning that almost one percent of individuals catching it eventually die from the disease. Only 67% of cases are symptomatic, the duration of sickness in mild symptomatic cases is 5 days and during this period individuals do not work. Approximately 1.3% of cases are severe enough to require critical care for 10 days on average. One-half of patients in critical care eventually dies and 0.023% of patients not requiring critical care also dies. All patients in a need of critical care die without it.

The calculation of new infections for laissez-faire and herd immunity approach corresponds to the predictions made for the US in Ferguson et al (2020), Figure A1. In the laissez-faire approach, the disease would end up infecting about 80% of the US population. The peak of the epidemic would happen in June with approximately 50% infected in some moment during that month. In the herd immunity approach mitigation and suppression measure would hold the number of case at the existing level of critical care capacity, which is around 138 th cases per month assuming that each critical care patient spends 10 days in an ICU. The epidemic and (most) mitigation measures would need to last until May 2022 to achieve the necessary herd immunity of 80% of population.

The calculation for the aggressive approach assumes that new cases stabilize in April and begin to decrease afterwards at the rate of 80% per month. After reaching 800 cases per month, the infection rate stabilizes due to missed and imported cases. We believe that this is a conservative estimate with respect to efficiency of aggressive mitigation measures. For comparison, in China the number of new cases went down from the peak of 3073 of new cases reported on February 11th to only 31 new case reported on March 11 (99% per month)^2^. In Italy, the number of new cases has been decreasing for three consecutive days since March 20th at the rate of at least 12% per day which corresponds to 98% decrease in one month.

### 2.3 Economic Costs

Economic costs come from three sources: direct costs of illness and treatment, employment losses and losses due to falling demand or mandatory shutdowns. This approach is consistent with the standard approaches used to estimate economic costs of the epidemic (Jonung and Roeger, 2006; Keogh-Brown et al, 2010).

We calculate direct costs of illness as costs of critical care stay for patients receiving critical care. The average cost per in-patient stay in the US is around 2000 USD, but varies across states and hospital types^3^. We assume that each day in an ICU increases hospital’s costs by 2000 USD and hence the average stay costs $20000 per each critical care patient with COVID-19.

Falling demand and mandatory shutdowns affect transport, food services, accommodation, arts and recreation. In the herd immunity approach, food services, arts and recreation shut down completely. Air transport loses 70% of output (note that about 15% of revenue in the US comes from mail and cargo) while ground transport and accommodation lose 50% of output for the duration of NPI measures (until May 2022). There are no mandatory shutdowns in the laissez-faire approach, but for peak infection month people avoid shopping in public places. For simplicity, we assume that the decrease in demand during the peak month of the epidemic (June 2020) is equal to the decrease in demand experienced due to social distancing measures in the herd immunity approach. In contrast to some older studies (Smith et al, 2009) we do not assume that the epidemic causes a decrease in the demand for goods given a widespread use of online shopping in the US.

The decrease in value added in affected sectors adds up to 4.4% of GDP. In our calculation, this is the total negative effect on GDP due to demand shocks and shut-downs. This estimate ignores the potential substitution from affected sectors to other goods and services. Accounting for substitution would make the negative effect smaller. On another hand, it also ignores spillovers effects to other sectors supplying goods and services, which could increase the actual effect. This negative effect on GDP lasts from April 2020 to May 2022 in the herd immunity approach, but happens only in June 2020 in the laissez-faire approach. These demand shocks also make redundant almost 8% of the US workforce.

Next, we consider the labor supply effects on output. The large part of output effects comes from employment losses due to sickness, home quarantines and school closures. Sickness losses equal to the number of symptomatic mild cases multiplied by 5 days (duration of symptoms) plus the number of critical cases multiplied by one month (average duration of critical cases). Total employment losses due to sickness are approximately equal for the laissez-faire approach and for the herd immunity approach as in both approaches the virus ends up infecting around 80% of population.

Home quarantines imposed in the herd immunity approach involve 14-day shelter-in-place orders for all the household members if one household members gets diagnosed with COVID-19. Consistent with Ferguson et al (2020), we assume that only 50% of household comply with quarantine orders. The average number of workers per household weighted by household size represents the expected number of workers affected by the household quarantine per each diagnosed case. Based on the American Community Survey 2018 obtained from I-PUMS^4^, this number is 1.15. Excluding workers which already quarantine due to sickness, the additional loss of labor supply is 14 *\** (1.15 *\** (1 *−s*) + (1.15 *−* 1) *\* s*) days, where *s* is the proportion of workers in the population (41% in the US).

School closures decrease labor demand by requiring that at least one adult needs to be present at home during school hours if the household includes any schoolchildren younger than 12 years old. To find all the affected households, we consider households in which the number of 18-80 individuals is equal to the number of full-time workers commuting to work and which have at least one child with age between 6 and 12 years. In the ACS 2018, these criteria describe approximately 7mln households and correspond to 5.2% of full-time workers.

Both demand shock, sickness and other restrictive measures affect the labor supply concurrently which somewhat reduces the total effect. To calculate the total effect of sickness, home quarantines and school closures net of demand effects we assume that these factors act independently from each other except sickness and home quarantines. We use the following formula to calculate the net effects on employment:

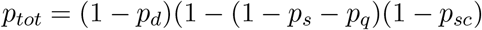

Where *p*_*tot*_ is the total effect on employment, *p*_*d*_ is the proportion of workers made redundant due to demand shocks and shutdowns, *p*_*s*_ is the proportion of workers falling sick, *p*_*q*_ is the proportion of labor supply lost due to HH quarantines and *p*_*sc*_ is the proportion of workers lost due to school shutdowns. The total effect on employment corresponds to the 6.6% decrease in employment during most of the epidemic.

In the laissez-faire approach, we assume that 33% of workers or all workers aged 50 and above do not show up for work during the peak epidemic month of June 2020 ^5^. This estimate is consistent with Sadique et al (2007) finding that 34% of workers expect to miss work in peak flu epidemic if mortality rates are high. The net effect on labor supply accounts for redundant workforce due to lower demand and sickness and corresponds to 36.2% less employment in June 2020.

We translate labor supply shocks into output by multiplying them by the average worker’s productivity in the US (GDP/Employment). This approach implicitly assumes zero elasticity of substitution between capital and labor^6^. This assumption is consistent with Grynza and Rycx (2018) who find in a matched employer-employee dataset of Danish firms that a one percentage point increase in sickness absenteeism reduces firm’s output by approximately one percent^7^.

As there is little guidance on potential productivity effects of most aggressive policies, we choose the most conservative pessimistic approach. We assume that the economy loses 100% of its labor force for the duration of these measures (3 months). This is the most conservative estimate given that some essential sectors still operate even in the case of Hubei province in China^8^.

Our preliminary estimates of testing and case tracking costs needed in the aggressive approach are negligible compared to costs of lost productivity and costs of mortality. Assuming that we test all residents experiencing common cold or flu symptoms which happens about 3 times per year (Arroll, 2011) and using the Medicare reimbursement rate of $51 as a measure of direct testing cost, the total testing costs of testing add up to approximately only $4 bln per month. Case tracking costs would be even smaller as long as infection rate remain in the predicted range. For example, hiring additional 1000 specialists to track cases with wages of $100,000 per year would cost less than $10 mln per month.

### 2.4 Mortality Costs

The population willingness to trade an increased risk of death for money or value of statistical life is the most crucial parameter of this cost-benefit analysis. There is a large uncertainty on the appropriate number for the value of statistical life (VSL). Two most common approaches to estimate VSL use either wage premiums in riskier occupations (compensating wage differentials approach) or directly ask for people’s willingness to pay for public programs to reduce mortality risks (stated preference approach). Viscusi and Aldi (2003) recommend the range of $7-12.4mln per life based on the meta-study of compensating wage differentials papers. Stated preference literature typically finds somewhat lower estimates. For example, the OECD recommends using the value of statistical life range of $1.8-5.5 mln based on the meta-study of stated preference literature.

To account for this uncertainty, we calculate the total costs of each program for two values of statistical life at opposite points of the plausible range. The low value of statistical life in our analysis is $3mln. The high value of statistical life is $10mln. For comparison, the US Environmental Protection Agency uses the value of statistical life of around $8.6mln (based on Alberini et al, 2019). There are indications that people are willing to pay similar amounts to save themselves from risks of death from SARS respiratory illnesses (Liu et al, 2005).

The literature finds that the value of statistical life slightly decreases with age for older people. The effect is much smaller as would be expected just based on expected years to live. Given the small size of this effect and the potential moral dilemmas, we avoid accounting for age variation in the VSL.

Mortality costs of the disease are highest for the laissez-faire approach in which the demand for critical care exceeds healthcare capacity starting from May 2020. It results in a large number of excess deaths: 3.5 mln vs 2.4 mln in the herd immunity approach. Accordingly, total mortality costs of the laissez-faire approach add up to $10.6 trillion for low VSL and $35.47 trillion for high VSL. Total mortality costs of the herd immunity approach are accordingly 7.1 and 23.8 trillion USD. Total mortality costs are almost negligible in the aggressive approach ($6.4bln and $21bln).

## 3 Results

Before proceeding to the discussion of the total costs, we would like to point out their crucial determinants. In terms of mortality, the laissez-faire approach results in higher costs followed by the herd immunity approach and by the aggressive approach. The order is exactly opposite for the costs on output of goods and services. Hence it is natural to expect that the the aggressive approach would achieve lower costs if we put high value lives saved or if the infection fatality rate is low, while the laissez-faire approach would be the least costly one if the value of statistical life is low or if the mortality is low.

Somewhat surprisingly, we find that the aggressive approach achieves the lowest costs for the society both for low and high values of statistical life. The total costs ($5500 bln) are more than 50% lower than the costs of the next best approach for low VSL and more than 80% for high VSL. The costs of shutting down the economy for 3 months are still lower than the costs of additional mortality experienced both in the herd immunity and in the laissez-faire approaches.

If the aggressive approach is unfeasible, the laissez-faire and herd immunity approaches result in similar total costs if using the low VSL, but the herd immunity approach achieves significantly lower costs for higher VSL (20% lower if VSL=$10mln). Given that the existing research tends to point to higher willingness–to-pay to avoid SARS infections and a higher estimate for VSL used by the Environmental Protection Agency, economic costs of suppression measures seems to be justified. However, given the relatively close estimate of total costs for both approaches, it would be interesting to see how these estimate would change if accounting for additional factors (see extensions) or by more accurate economic projections.

Note, that both the aggressive approach and the herd immunity approach have lower costs despite quite pessimistic assumptions applied in these scenarios. For example, the aggressive approach takes longer than the lockdown in Hubei province of China which is finishing in two months and does not completely stop the economy. For the herd immunity approach, we assume no healthcare capacity expansion within 2 years of the epidemic. And the laissez-faire approach assumes that people’s precautionary responses are limited to one month only (June).

How low should be the disease mortality rate to justify using the laissez-faire approach? Our calculations show that for the infection fatality rate of 0.15% and lower doing nothing would minimize total societal costs for the whole range of value of statistical life ($3-10 mln). For comparison, 2009 swine flu had a mortality rate of 0.025% in the UK (Donaldson et al, 2020) and lower attack rates. Seasonal flu has a slightly higher mortality, but also lower attack rates.

**Table 1:** Total Inclusive Costs of Mitigation Approaches

| Approach | Total societal costs (\$bln) | |
| --- | --- | --- |
|  | Minimum value of life | Maximum value of life |
| Laissez-faire | 11,469 | 35,470 |
| Herd immunity | 11,524 | 27,527 |
| Aggressive | 5,439 | 5,454 |

## 4 Limitations and Possible Extensions

### Increasing Healthcare Capacity

Increasing critical care capacity would allow to infect a larger number of individuals each month. This would have no effect on the total mortality, but would decrease the duration of suppression measures and reduce their economic impact. Accounting for this factor would decrees total costs of the herd immunity approach.

### Gradual Removal of Suppression Measures in the Herd Immunity Approach

As the proportion of susceptible population goes down, the basic reproduction number *R*_0_ also goes down. It follows from the formula *R*_0_ = *βsγ* where *β* is the number of contacts made by each person and *s* is the proportion of susceptible individuals among these contacts. It means that in the herd immunity approach there would be a possibility and a need to stop some NPI (school closures?) to keep the number of new infection at the capacity and to reduce economic costs. Accounting for these changes would decrease total costs of the herd immunity approach.

### Duration of Immunity

The current calculation assumes that the recovering from the disease grants lasting immunity to it. This is not the case for many other coronaviruses causing common cold (Callow et al, 1993) which give immunity for about one year. If the immunity is going to last less than two years, the herd immunity approach would never end the epidemic on its own. Each month the susceptible population would increase as individuals catching disease several months ago would lose immunity.

### Complete Virus Eradication

Laissez-faire approach would infect more than 70% of population in less than in 4 months. Assuming that recovering from the disease grants immunity for at least one year, the population would achieve herd immunity and the epidemic would stop. It means that there is a significant probability that the virus would completely disappear in the laissez-faire approach as it happened with the virus responsible for the 1918 flu pandemic. More epidemiological analysis and expertise is needed to estimate the probability of this event.

### Decreasing Flu and Common Cold Attack Rates

Non-pharmaceutical measures considered in this analysis also decrease the transmission of other airborne infectious diseases, including flu, common cold, pertussis and measles. For exampe, during the 2019-2020 season flu has infected around 36 mln people in the US and caused 370 thousand hospitalizations and 22 thousand deaths. The average annual monetary cost of seasonal flu epidemics is $87bln in 2003 prices (Molinari et al, 2007) or $163bln if accounting for higher population, prices and productivity in 2019. Fendrick et al (2003) estimate the total economic costs of common cold for the US at around $40 bln in 2001 prices (or $81bln accounting for price/population/productivity growth to 2019). These benefits extend mostly to the herd immunity approach and partially to the agressive approach.

### Leisure

All the approaches increase the amount of leisure to a different degree. While leisure is valuable to most people, the existing empirical research gives little guidance on monetary value of leisure with social distancing.

### Possibility to Develop a Treatment/Vaccine

The development of a vaccine or successful treatment would reduce the mortality rate of the disease and potentially decrease its attack rate. A higher probability of treatment or vaccine would reduce total costs of the herd approach by limiting its duration. We consider this outcome to be unlikely given the absence of vaccines or effective treatments for other coronaviruses.

### Incomplete Risk Sharing

If an arbitrary reallocation of money is possible between different individuals, then the approach with lowest total costs can make everybody better off ex ante (before knowing personal survival outcome). However, no existing plans consider wide-scale redistribution. In this conditions, other measures can give more accurate description of welfare gains and losses, such as ex ante expected utility in a heterogeneous agent model.

## Data Availability

All the data used to prepare this preprint is publicly available. The calculation is available as a supplementary material.

https://drive.google.com/file/d/1V5j8lKTIOAaLhGQJCJKsqlwGHse1MHrQ/view?usp=sharing

---

**Table 2:** Demand Shocks

| Sector | Share |  | Relative | Shock |  |
| --- | --- | --- | --- | --- | --- |
|  | GDP | Empl. |  | GDP | Employment |
| **Agriculture, forestry, –** | 0.8 |  | 0 | 0 | 0 |
| **Mining** | 1.4 |  | 0 | 0 | 0 |
| **Utilities** | 1.6 |  | 0 | 0 | 0 |
| **Construction** | 4.1 |  | 0 | 0 | 0 |
| **Manufacturing** | 11 |  | 0 | 0 | 0 |
| **Wholesale trade** | 6 |  | 0 | 0 | 0 |
| **Retail trade** |  |  |  |  | 0 |
| Motor vehicle and parts dealers | 1 |  | 0 | 0 | 0 |
| Food and beverage stores | 0.8 |  | 0 | 0 | 0 |
| General merchandise stores | 0.7 |  | 0 | 0 | 0 |
| Other retail | 3 |  | 0 | 0 | 0 |
| **Transportation and warehousing** |  |  |  | 0 | 0 |
| Air transportation | 0.7 | 0.43 | 0.7 | 0.49 | 0.301 |
| Rail transportation | 0.2 |  | 0 | 0 | 0 |
| Water transportation | 0.1 |  | 0 | 0 | 0 |
| Truck transportation | 0.8 |  | 0 | 0 | 0 |
| Transit and ground passenger | 0.2 | 0.68 | 0.5 | 0.1 | 0.34 |
| Pipeline transportation | 0.2 |  | 0 | 0 | 0 |
| Other transportation | 0.6 |  | 0 | 0 | 0 |
| Warehousing and storage | 0.4 |  | 0 | 0 | 0 |
| **Information** | 5.2 |  | 0 | 0 | 0 |
| **Finance, insurance, –** | 20.9 |  | 0 | 0 | 0 |
| **Professional, – services** | 7.7 |  | 0 | 0 | 0 |
| **Management of companies –** | 2 |  | 0 | 0 | 0 |
| **Administrative services** | 3.2 |  | 0 | 0 | 0 |
| **Educational services** | 1.2 |  | 0 | 0 | 0 |
| **Health care** |  |  |  | 0 | 0 |
| Ambulatory health care | 3.7 |  | 0 | 0 | 0 |
| Hospitals | 2.4 |  | 0 | 0 | 0 |
| Nursing and residential care | 0.7 |  | 0 | 0 | 0 |
| Social assistance | 0.7 |  | 0 | 0 | 0 |
| **Arts, entertainment, and recreation** | 1.1 | 1.66 | 1 | 1.1 | 1.66 |
| **Accommodation and food services** |  |  |  |  | 0 |
| Accommodation | 0.8 | 1.03 | 0.5 | 0.4 | 0.515 |
| Food services and drinking places | 2.3 | 4.9 | 1 | 2.3 | 4.9 |
| **Other services, except government** | 2.1 |  | 0 | 0 | 0 |
| **Government** | 12.3 |  | 0 | 0 | 0 |
| **Total** | 100 | 100 | - | 4.4% | 7.7% |

## Footnotes

1. Reuters based on Goldman Sachs report https://www.reuters.com/article/us-health-coronavirus-china-toll/goldman-sees-chinas-economy-shrinking-9-in-first-quarter-amid-coronavirus-outbreak-idUSKBN21340T

2. WHO situation reports.

3. https://www.beckershospitalreview.com/finance/average-cost-per-inpatient-day-across-50-states.html

4. Steven Ruggles, Sarah Flood, Ronald Goeken, Josiah Grover, Erin Meyer, Jose Pacas and Matthew Sobek. IPUMS USA: Version 10.0 [dataset]. Minneapolis, MN: IPUMS, 2020. https://doi.org/10.18128/D010.V10.0

5. Working in June increases chances to catch the disease for non-immune individuals by approximately 20% with additional daily risk of around 0.007%. Given the risk of death conditional on disease of 0.6% for the group of 50-60 yrs old, the daily mortality risk outweighs the wages lost due to absenteeism (around $170 per day in the US) for most workers assuming the value of statistical life of $3mln.

6. The alternative approach would be to use the more flexible production function (such as Cobb-Douglas) to account for substitution which would reduce projected output losses.

7. Note also that if we take the predicted labor supply effects in more sophisticated general equilibrium models (as Smith et al, 2009 and Keogh-Brown et al, 2010) and take them as predictions of output effects, the resulting output effects would closely match the predictions of general equilibrium models (within 20%, not percentage points!) as their GE multiplier effects compensate lower elasticities of substitution between capital and labor.

8. Including at least grocery stores, pharmacies, healthcare (National Geographic) https://www.nationalgeographic.com/history/2020/03/60-million-person-coronavirus-lockdown-united-states/

